# Seroprevalence of anti-SARS-CoV-2 antibodies in COVID-19 patients and healthy volunteers

**DOI:** 10.1101/2020.08.30.20184309

**Authors:** Patrícia Figueiredo-Campos, Birte Blankenhaus, Catarina Mota, Andreia Gomes, Marta Serrano, Silvia Ariotti, Catarina Costa, Helena Nunes-Cabaço, António M. Mendes, Pedro Gaspar, M. Conceição Pereira-Santos, Fabiana Rodrigues, Jorge Condeço, M. Antonia Escoval, Matilde Santos, Mario Ramirez, José Melo-Cristino, J. Pedro Simas, Eugenia Vasconcelos, Ângela Afonso, Marc Veldhoen

**Author notes:** Shared first authors (Patrícia Figueiredo-Campos, Birte Blankenhaus). Correspondence to M.V., - ext: 47250.

## Abstract

SARS-CoV-2 has emerged as a novel human pathogen, causing clinical signs, from fever to pneumonia – COVID-19 – but may remain mild or even asymptomatic. To understand the continuing spread of the virus, to detect those who are and were infected, and to follow the immune response longitudinally, reliable and robust assays for SARS-CoV-2 detection and immunological monitoring are needed and have been setup around the world. We quantified immunoglobulin M (IgM), IgG and IgA antibodies recognizing the SARS-CoV-2 receptor-binding domain (RBD) or the Spike (S) protein over a period of five months following COVID-19 disease onset or in previously SARS-CoV-2 PCR-positive volunteers. We report the detailed setup to monitor the humoral immune response from over 300 COVID-19 hospital patients and healthcare workers, 2500 University staff and 187 post-COVID19 volunteers, and assessing titres for IgM, IgG and IgA. Anti-SARS-CoV-2 antibody responses followed a classic pattern with a rapid increase within the first three weeks after symptoms. Although titres reduce from approximately four weeks, the ability to detect SARS-CoV-2 antibodies remained robust for five months in a large proportion of previously virus-positive screened subjects. Our work provides detailed information for the assays used, facilitating further and longitudinal analysis of protective immunity to SARS-CoV-2. Moreover, it highlights a continued level of circulating neutralising antibodies in most people with confirmed SARS-CoV-2, at least up to five months after infection.

## Introduction

SARS-CoV-2 infection, causes a wide variety of disease symptoms, from fever, asthenia or myalgia, to pneumonia and in most severe cases acute respiratory distress syndrome, referred to as COVID-19. Yet, a large amount of SARS-CoV-2 infected patients remains asymptomatic. SARS-CoV-2 rapid spread around the world was declared a global pandemic in March 2020. It remains a continuing threat to health and socio-economic wellbeing. Despite the global number of infections reaching tens of millions, including almost one million fatalities, due to mitigation measures, the overall infection rate is relatively low with local infection hotspots. Although scientific progress is rapid, there remains a pressing need to understand the immune response that follows SARS-CoV-2 infection, including its role during disease and especially its potential long-term protective effects.

A prime immune target during coronavirus infections is the spike (S) protein, closely associated with and targeted by neutralising antibody responses and protective immunity, in contrast to most other viral proteins [1 -4]. The S protein is responsible for the interaction of SARS-CoV-2 with the host cells via binding ACE2 [5-7]. It can be divided into two regions, S1 and S2. The extra-viral S1 region contains within its second domain the receptor binding domain (RBD) [8]. The SARS-CoV-2 RBD sequence shows limited homology with seasonal coronaviruses or EMC/2012, the cause of Middle East respiratory syndrome (MERS). In contrast, SARS-CoV-2 RBD shares 73% of its sequence with the RBD of SARS [3].

Attempts to curtail and control the SARS-CoV-2 virus rely on increasing inter-personal distance, including the closure of much social and economic activity, as well as testing for acute infection and personal hygiene measures. This was implemented early during the outbreak, with the University of Lisbon closing after March 13^th^, 10 days after the first recorded cases in Portugal. However, during the subsequent transition phase, restrictions have steadily been lifted. The gradual return to social and economic activity requires active surveillance to determine local outbreaks, contact tracing and quarantine. In addition, those most vulnerable to COVID-19 will need to remain under enhanced protection. Important information is how protective immunity develops in the population at large and in specific groups such as healthcare professionals. A thorough assessment of the duration of protective immunity is critical to determine the measures that need to be taken to prevent and handle future waves of SARS-CoV-2. Such information will need to be gathered widely, in different locations around the world, reflecting local conditions, such as containment measures and their timing. The data obtained will need to be accurate and the methods used transparent and reproducible to enable comparisons between locations and countries. The recent SARS-CoV-2 outbreak brings limitations with respect to exposure time, but also gives us the opportunity to acquire real-time data and develop reliable longitudinal follow up studies.

To determine the cumulative rate of infection in communities and gaining insight into the potential protection against re-infection, serological assays are critical. Depending on the aims of the study, the setup of such assays can be used for the detection of exposure to SARS-CoV-2 as well as gaining insights into neutralisation activity, since antibody titres for both the S protein and RBD have been shown to correlate well with neutralising activity [3, 9-11]. We describe the detailed setup and versatility of a seroconversion assay to determine humoral immunity to SARS-CoV-2 that was used for screening hospital patients, healthy post-COVID19 volunteers and staff of the University of Lisbon. We report that in the acute phase men produce more antibodies than women, but levels equilibrate during the resolution phase and are similar between the genders in the months after SARS-CoV-2 infection. We show that antibodies against SARS-CoV-2 Spike and its RBD domain are readily detectable in the majority of cases, including in patients receiving immune suppressive or anti-retroviral therapy. In line with a classic immune response, SARS-CoV-2 antibodies in the blood peak around week 3 post infection, and although titres reduce, IgG antibodies remain detectable for at least 5 months.

## Results

### Seroconversion assay setup

To detect seroconversion, point-of-care devices are practical and desirable. However, without the investment in development and the proper equipment, the use of lateral flow assays have limited success with respect to sensitivity and antibody titres cannot be determined. The gold standard for antibody detection remains the enzyme-linked immunosorbent assay (ELISA), offering high flexibility and sensitivity, but limited scalability [11]. SARS-CoV-2 Spike is a prominent immunogenic antigen and its RBD is least conserved compared with other coronaviruses. Hence, the use of Spike and its RBD quickly became the focus of seroconversion assays. We chose for the present study the assay developed by Florian Krammer and his laboratory, a format that received FDA emergency approval in April 2020 and is described in detail [12].

Human sera pose a biological hazard to laboratory workers and can potentially contain not only SARS-CoV-2, but also other infectious viruses. Therefore, all ELISA steps were performed at biosafety level (BSL) 2, with BSL3 personal protective equipment. Inactivation procedures are recommended but can have uncertain effects on the accuracy of serological testing [13]. We tested three common procedures: a) one hour heat inactivation at 56°C, or b) the addition of a non-ionic surfactant (0.1% Triton X-100), or c) the combination of both, in comparison to neat serum. Serial dilutions of two chosen SARS-CoV-2 PCR-positive serum samples showed IgG detection following all three inactivation methods and was indiscriminate from untreated controls (Fig.1A).

**Figure 1.**
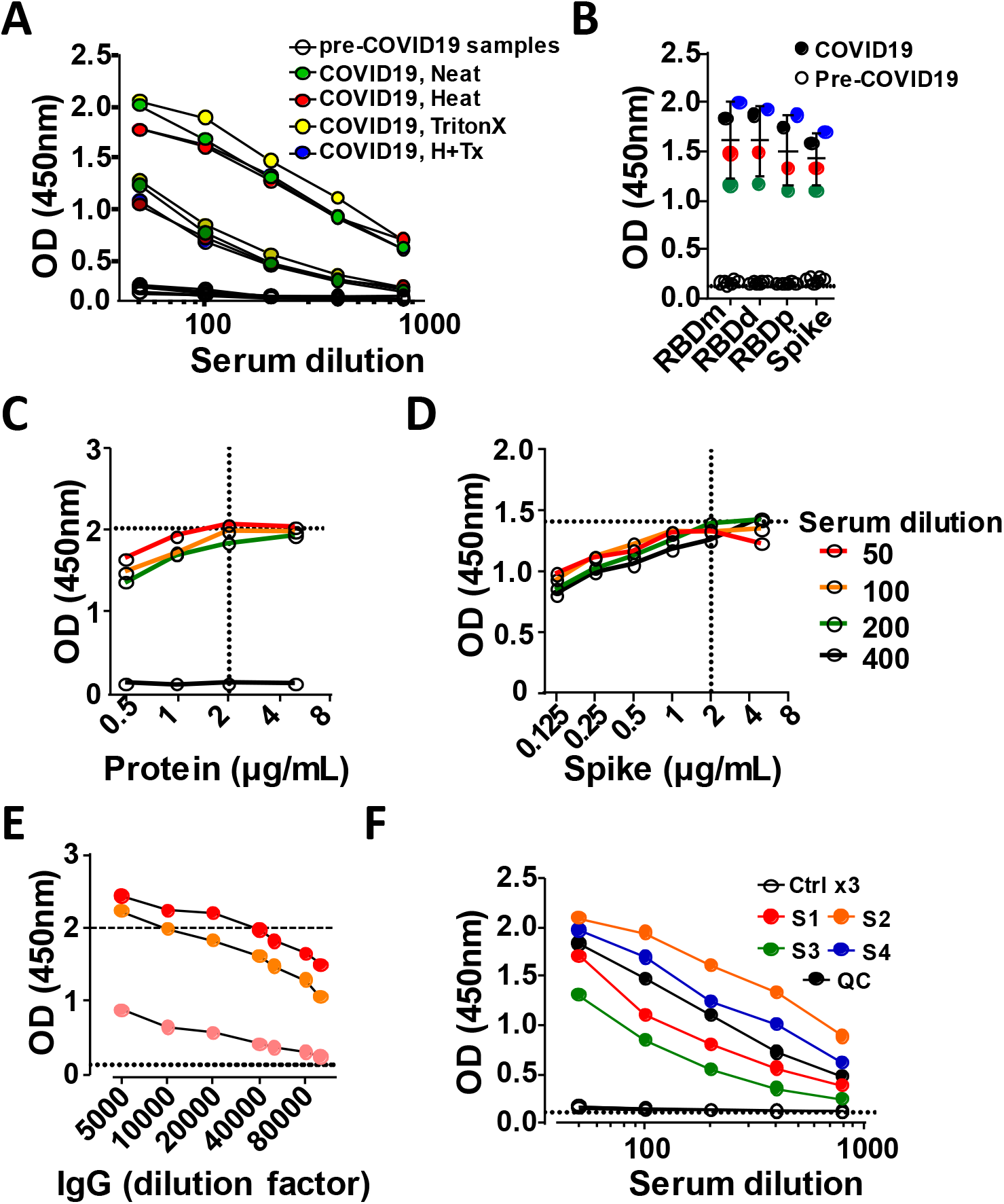
SARS-CoV-2 ELISA setup. SARS-CoV-2 IgG antibody detection in serum samples from SARS-CoV-2 PCR-positive subjects or pre-COVID-19 controls using Immulon 4HBX 96-well plates coated with RBD protein. Absorbance (Optical density-OD) was evaluated at 450 nm. **A**) Three serum samples, pre-COVID-19 control, medium and high titre were treated by the indicated methods to inactivate virus particles. **B**) Isolated RBD monomer, RBD dimer, pooled RBD and Spike protein were used for coating at the same conditions and 4 COVID-19 serum samples (coloured) were tested versus four pre-COVID-19 (open symbols). **C-D**) 96-well plate was coated with C) RBD or D) Spike protein at indicated concentrations and three COVID-19 (coloured) and pre-COVID-19 (black) sera were tested. **E**) Secondary antibody dilution titration anti-IgG, at indicated dilution on 96-well plate coated with 2 μg/ml RBD protein. **F**) Quality control (QC) and sample serum serial dilution. QC sample is a mix of the serum samples tested from four healthcare workers (S1-S4). Dashed line indicates blank values.

The S protein has a trimeric structure, while the *in vitro* expression of RBD results in the generation of monomeric and dimeric protein. However, when we tested the ability of RBD mono- and dimeric protein for antibody binding, both performed similar and comparable to the total protein fraction (Fig.1 B). Additional parameters affecting the performance of ELISA assays, such as the coating time (o/n – 1 week at 4°C), serum incubation time and temperature, as well as the amount of tetramethylbenzidine (TMB) substrate and the development time (adjusted to 10 minutes) were optimised (Suppl.Fig.1A, data not shown). Coated plates were stable for a week, and incubation of 1 or 2 hours at room temperature or 37°C were indistinguishable. To ensure that the ELISAs runs at non-saturating conditions, we performed a full titration of the capture antigens (from 0.125 μg/mL-10μg/mL) and the secondary antibodies (1:5000-1:100000) used for antibody detection. SARS-CoV-2 proteins were titrated and assessed using individual patient sera (high, medium and low titres). Simultaneously, the secondary antibodies were titrated, such as anti-IgG, but also anti-IgM, anti-IgA and anti-total Ig, whereby optical density (OD) of 2.0 was used as an upper limit to avoid saturation of the assay (Fig.1C-E, Suppl.Fig.1B-G).

In order to prepare for diagnostic use, the generation of quality control (QC) serum is critical to validate each assay run. Sera from exposed patients is most desirable as it will contain antibodies with a range of avidities and isotypes, providing more stable binding properties. However, sufficient volume needs to be obtained to ensure that there is enough material to complete the ELISA validation process and the study or series of studies to be undertaken. Sera from four SARS-CoV-2 exposed but healthy volunteers were assessed and pooled to serve as quality control for subsequent assays (Fig.1 F). Ultimately, antibody signals should diminish in a dose-dependent way using serial dilutions of the sera, enabling the accurate determination of antibody titres (Suppl.Fig.1H).

### Seroconversion assay validation

We used 100 pre-COVID-19 sera from healthy volunteers collected between October 2012 and November 2017 as negative controls (Table 1). Furthermore, we obtained 19 sera from PCR positive hospital healthcare workers with mainly mild symptoms, just over 30 days since first symptoms and the positive SARS-CoV-2 PCR result (Table 2). Seroconversion was detected in the sera of all SARS-CoV-2 PCR-positive patients using the RBD part of SARS-CoV-2 S antigen and 18/19 using the full-length SARS-CoV-2 S protein by probing for IgG (Fig.2A). Receiver operating characteristic curve (ROC) determined sensitivity and specificity and the assays cut-off, at 0.4171 and 0.4816 for RBD and S protein respectively, corresponding to 100% specificity and 99% sensitivity for RBD and 94.74% specificity and 98% sensitivity for Spike in this initial analysis (Fig.2B-C, Table 3).

**Figure 2.**
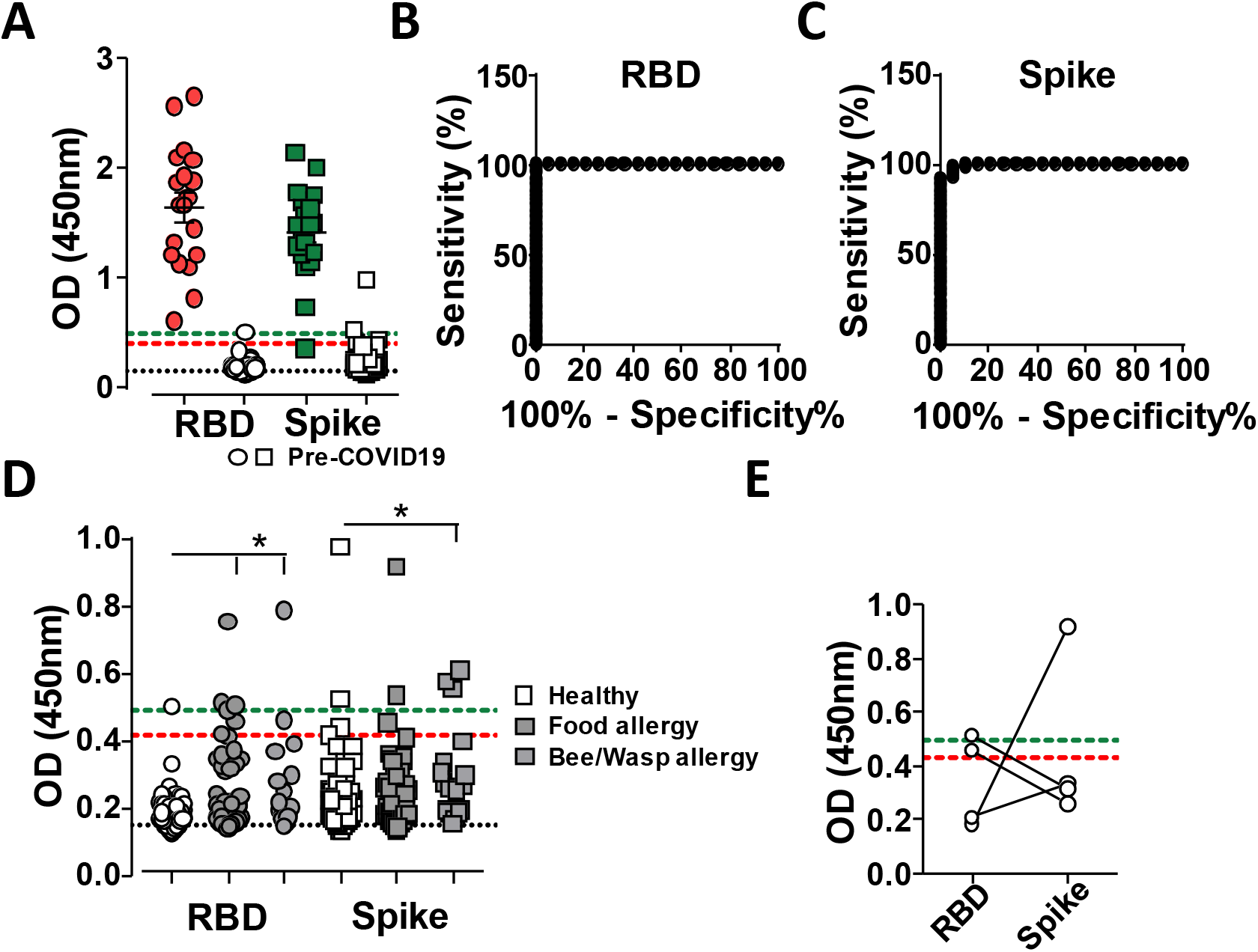
SARS-CoV-2 ELISA testing. SARS-CoV-2 IgG antibody detection in serum samples from SARS-CoV-2 PCR-positive subjects (coloured) or pre-COVID-19 controls (open) using Immulon 4HBX 96-well plates coated with RBD (circles) or Spike (squared) proteins. Absorbance was evaluated at 450 nm. **A**) Serum at 1 /50 dilution from 19 SARS-CoV-2 PCR-positive healthcare workers were assessed for anti-SARS-CoV-2 RBD and Spike IgG and compared with 100 pre-COVID-19 sera. **B-C**) ROC analysis, plotting sensitivity against specificity of B) RBD or C) Spike samples as shown in A). **D**) Serum at 1/50 dilution from pre-COVID-19 cohorts, healthy (100, open symbols), food allergies (62, dark grey symbols) and bee/wasp allergies (30, light grey symbols), were tested for RBD and Spike protein reactivity. **E**) Example of cross-reactivity on RBD or Spike protein from pre-COVID-19 serum as used in D). Statistical analysis was performed using Kruskal-Wallis test with Graphpad Prism software. * p<0.05, *** p<0.001. Dashed lines indicate, Black: blank values, Red: RBD cut off, Green: Spike cut off.

**Table 1.**
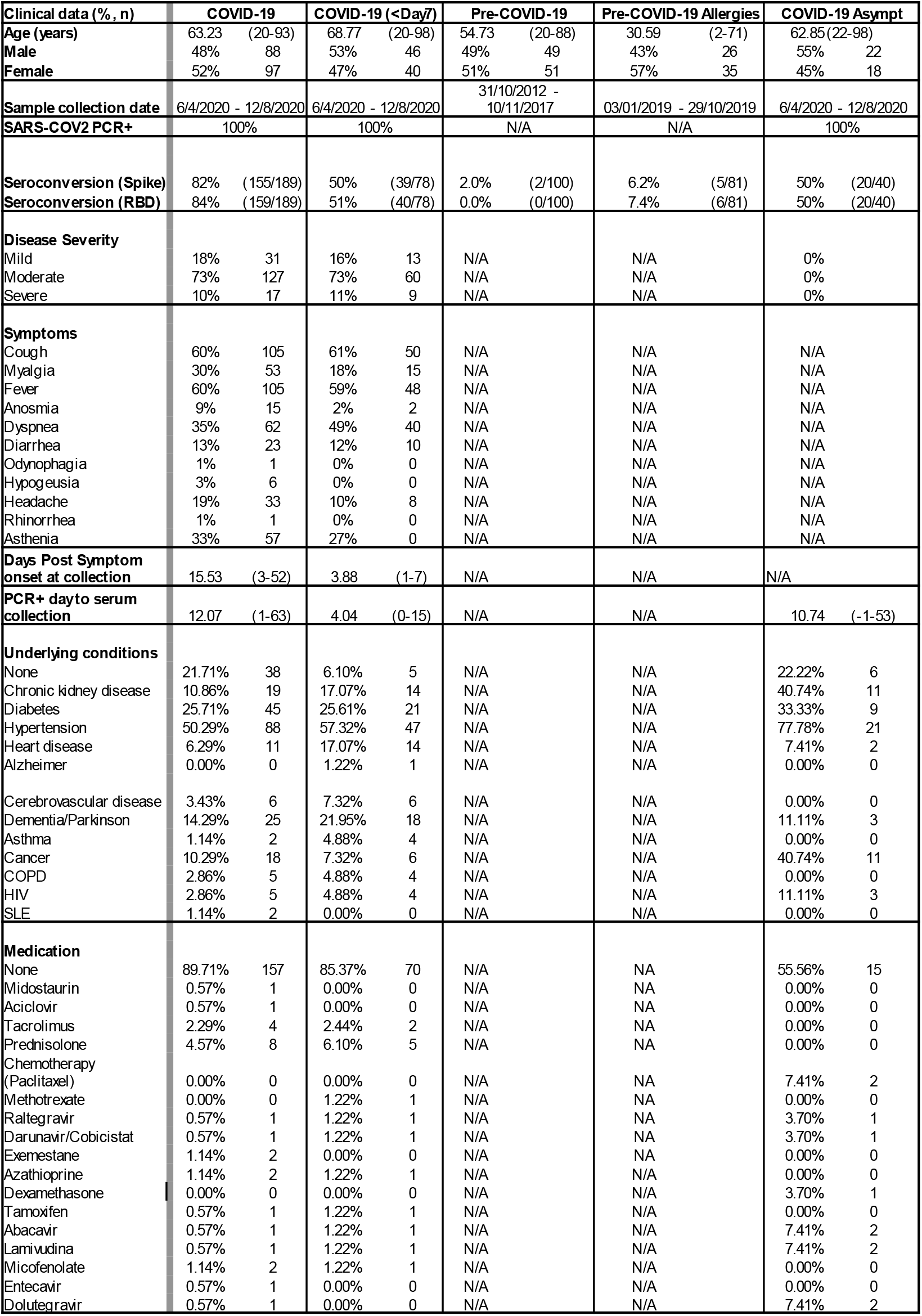
Demographics of patient participants, disease severity categories and symptoms, underlying conditions and medication.

**Table 2.**
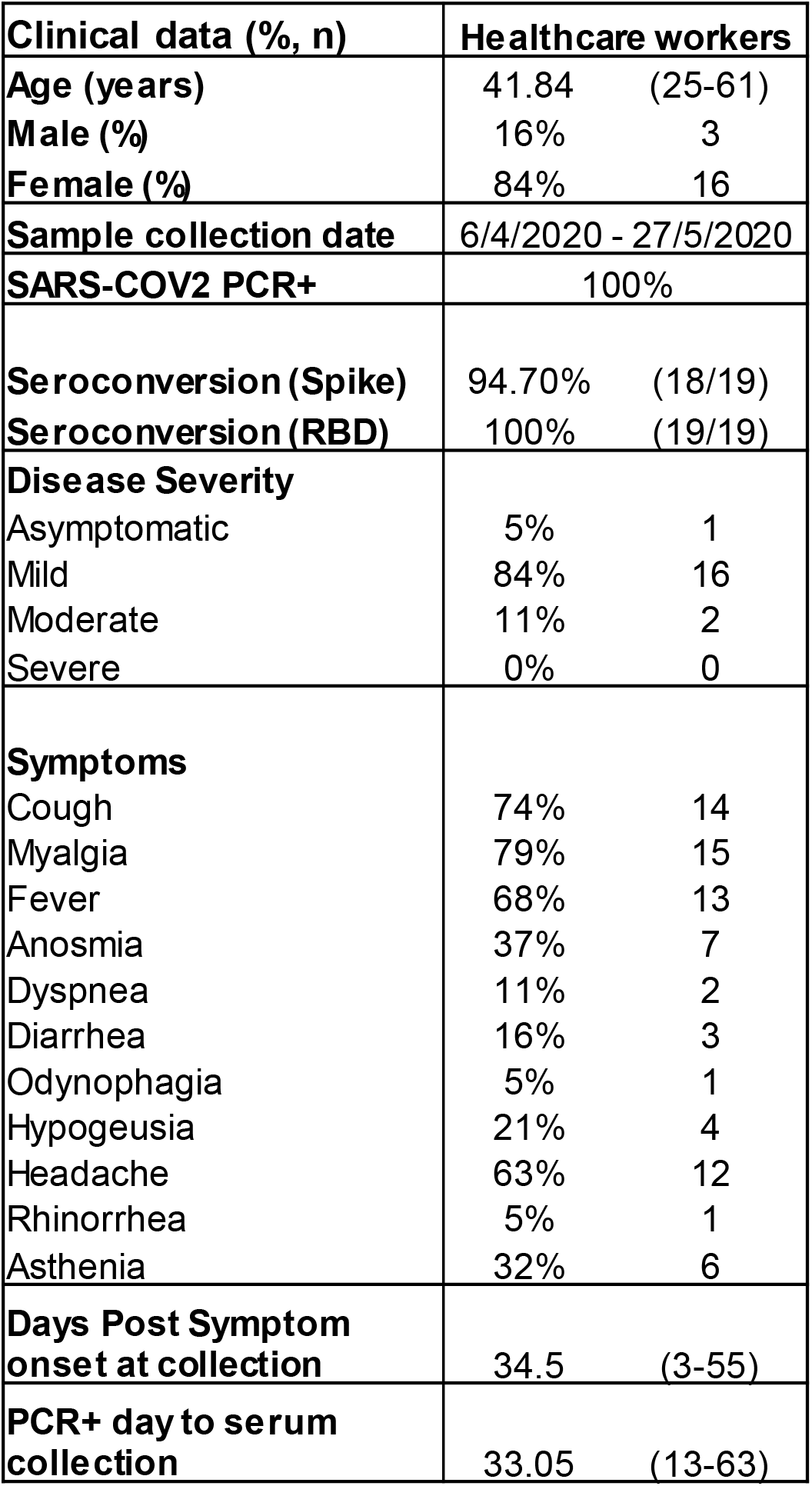
Demographics of healthcare participants, disease severity categories and symptoms.

| **Clinical data (% , n)** | **Healthcare workers** |  |
| --- | --- | --- |
| **Age (years)** | 41.84 | (25-61) |
| **Male (%)** | 16% | 3 |
| **Female (%)** | 84% | 16 |
| **Sample collection date** | 6/4/2020 - 27/5/2020 |  |
| **SARS-COV2 PCR+** | 100% |  |
| **Seroconversion (Spike)** | 94.70% | (18/19) |
| **Seroconversion (RBD)** | 100% | (19/19) |
| **Disease Severity** |  |  |
| Asymptomatic | 5% | 1 |
| Mild | 84% | 16 |
| Moderate | 11% | 2 |
| Severe | 0% | 0 |
| **Symptoms** |  |  |
| Cough | 74% | 14 |
| Myalgia | 79% | 15 |
| Fever | 68% | 13 |
| Anosmia | 37% | 7 |
| Dyspnea | 11% | 2 |
| Diarrhea | 16% | 3 |
| Odynophagia | 5% | 1 |
| Hypogeusia | 21% | 4 |
| Headache | 63% | 12 |
| Rhinorrhea | 5% | 1 |
| Asthenia | 32% | 6 |
| **Days Post Symptom onset at collection** | 34.5 | (3-55) |
| **PCR+ day to serum collection** | 33.05 | (13-63) |

In order to increase pre-COVID-19 sample size and reflect a broader spectrum of the population, we obtained 60 samples of individuals with food allergies and 30 samples from individuals with bee and wasp allergies, because these contain increased levels of antibodies [14]. Serum from allergic subjects increased the observed background on both RBD and Spike proteins (Fig.2D). Of importance, increased reactivity to one protein was often not observed on the second SARS-CoV-2 protein (Fig.2E), substantiating the two-step process of screening for RBD and subsequently those sera found positive for Spike [12].

### Seroconversion screening of COVID-19 hospitalised patients

We subsequently analysed 307 samples from hospitalised patients who tested positive for SARS-CoV-2 by PCR for the presence of antiSARS-CoV-2 IgG. Samples were acquired between April 6^th^ and August 12^th^, 2020 and at different times after the development of COVID-19 symptoms. The patients demonstrated a variety of symptoms and underlying medical conditions (Table 1). Seroconversion screening resulted in varied optical density measurements (Fig.3A). In line with the number of days normally taken by adaptive cellular immunity to be initiated, antibody detection in samples 14-days past first symptoms was robust in 78/78 (100%) samples on RBD and 76/78 (97.4%) on Spike protein (Fig.3B). Although antibody responses take time to mature, around half of the samples showed robust IgG seroconversion within the first week of symptoms, 40/78 (51 %) and 39/78 (50%) on RBD and Spike respectively (Fig.3C). Follow up samples from 68 patients showed that those who had seroconverted in the first week of symptoms maintained high levels of IgG one week later (27/27, 100%). From those who did not have an IgG response within the first week, 30/41 (73%) showed a robust response seven days later. The remaining 11 /41 (27%) patients had their second sample analysed in the second week after onset of symptoms, but still did not show an IgG response. However, although some patients did not show an IgG response in the second or even third week after onset of symptoms, those that were tested again one week later all seroconverted (10/10) (Fig.3D). From those who did not seroconvert within week two of COVID-19 symptoms (11/41, 27%), some had underlying conditions, such as systemic lupus erythematosus (SLE) (1), lymphoma (1), chemotherapy (1), or immunosuppressive medication (3).

**Figure 3.**
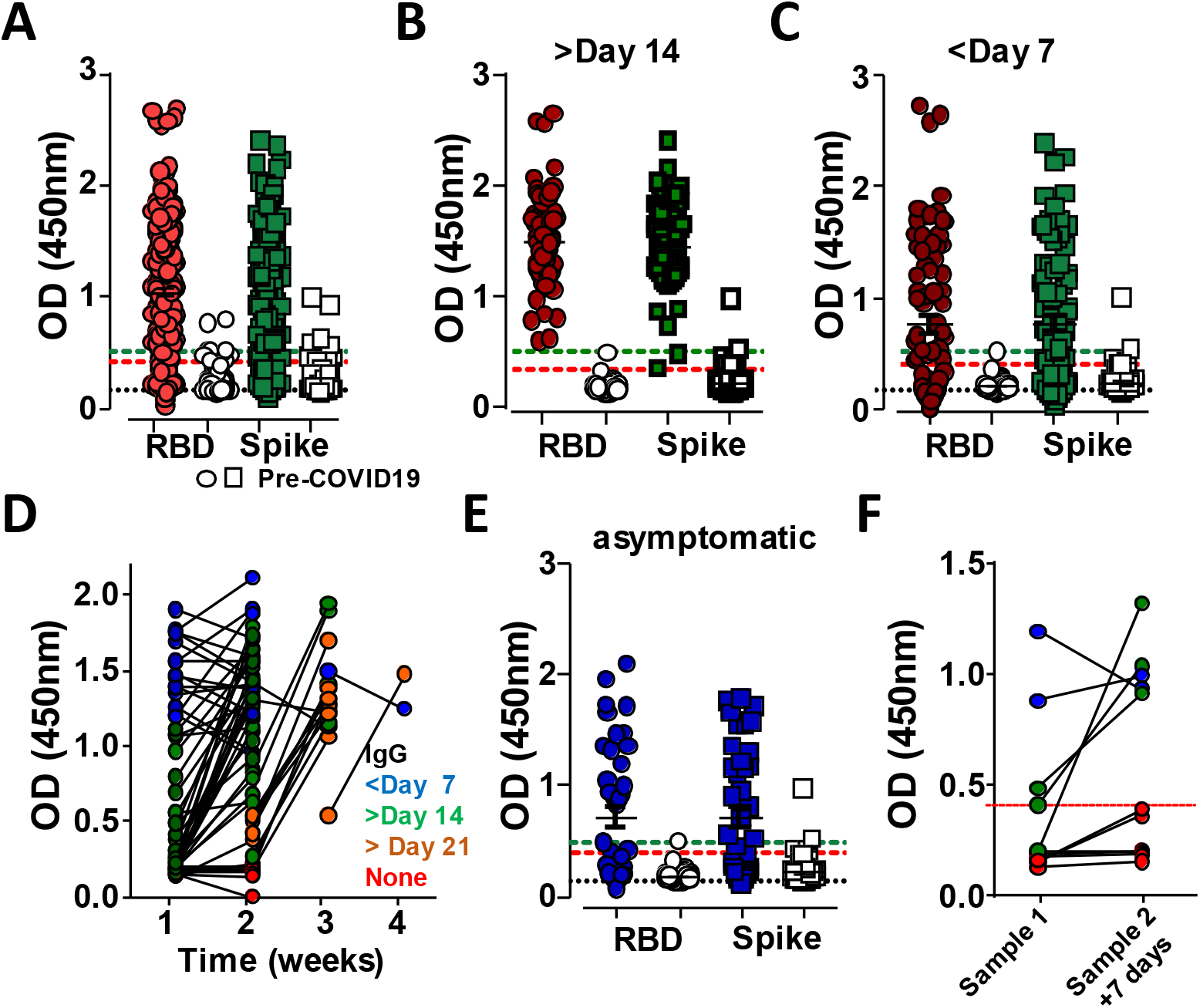
Seroconversion in hospitalised patients. SARS-CoV-2 IgG antibody detection in serum samples from SARS-CoV-2 PCR-positive hospital patients (coloured) or pre-COVID-19 controls (open) using Immulon 4HBX 96-well plates coated with RBD (circles) or Spike (squared) proteins. Absorbance was evaluated at 450 nm. **A**) Overview of over 300 SARS-CoV-2 PCR-positive tested subjects from Hospital Santa Maria and accumulated pre-COVID-19 controls. **B**) Selected samples presented in A) post-day 14 of initial reported COVID-19 symptoms (n=78). **C**) Selected samples presented in A) pre-day seven of the initial reported COVID-19 symptoms (n=78). **D**) Longitudinal follow up of patients sampled at the indicated week of COVID-19 symptom onset and re-sampling seven days later (n=76). Blue indicates continues high signal, Green those that seroconverted at the second sampling in week two, Orange those that seroconverted at the second sampling past week two, Red those in which no seroconversion was detected in first and second sampling. **E**) Selected samples presented in A) without reported COVID-19 symptoms (n=40). **F**) Longitudinal follow up of asymptomatic patients sampled in the first week of COVID-19 symptom onset and re-sampling seven days later, colours as used as in D) (n=10). Dashed lines indicate, Black: blank values, Red: RBD cut off, Green: Spike cut-off.

Some patients who were SARS-CoV-2 PCR positive within an average of 8.7 days (−1 - 53) after blood was taken, did not show any classic COVID-19 symptoms (Table 1). Of these, 20/40 (50%) showed seroconversion for anti-SARS-CoV-2 IgG (Fig.3E). Since no symptoms were reported, it remained unclear at what stage of the infection these patients were. Those who were IgG negative may have been within the first days of infection, or antibody levels were very low. However, repeat sampling from several patients seven days later revealed that only 3/11 (27%) patients seroconverted, although optical density remained modest, while in 6/11 (55%) anti-SARS-CoV-2 RBD IgG levels remained below the assay’s cut off (Fig.3F). This suggest a limited or much delayed seroconversion response.

### Effect of demographics, immunomodulatory and anti-viral medication on seroconversion

IgG seroconversion, 14-days after onset of symptoms, was detected equally well between female and male patients, independently of age, and for both RBD and spike protein (Fig.4A-B). Seroconversion detection or the antibody response, since patients were assayed on average 8.5 days after being SARS-CoV-2 PCR positive, were reduced or delayed in those asymptomatic for COVID-19 compared with those experiencing COVID-19 symptoms (Fig.4C). In line with an adaptive immune response taking several days to develop, the main factor influencing seroconversion was time since onset of COVID-19 symptoms (Fig.4D).

**Figure 4.**
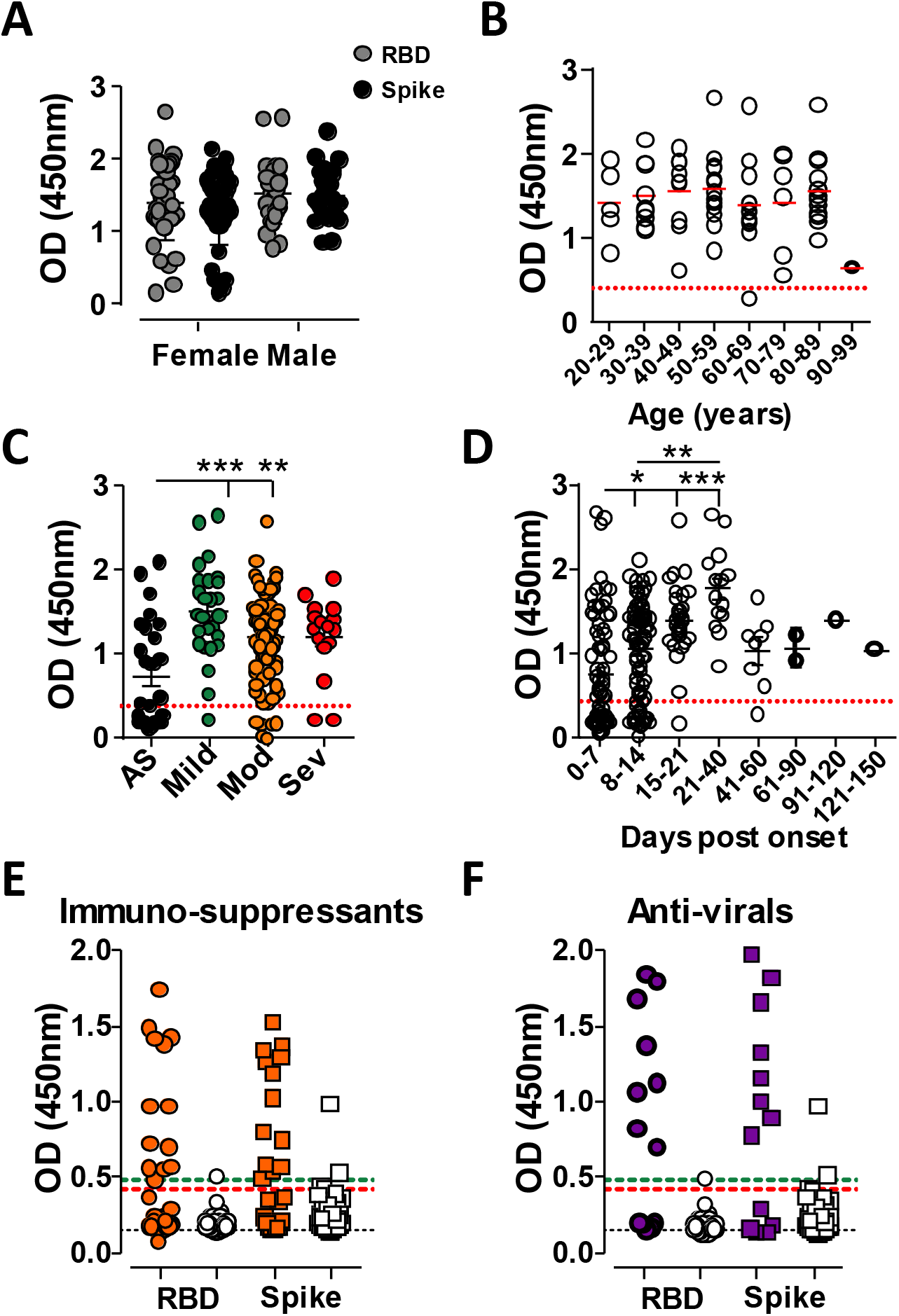
Seroconversion in subgroups and time. SARS-CoV-2 IgG antibody detection in serum samples from SARS-CoV-2 PCR-positive hospital patients or pre-COVID-19 controls using Immulon 4HBX 96-well plates coated with RBD (circles) or Spike (squared) proteins. Absorbance was evaluated at 450 nm. **A-B**) IgG OD signals were plotted of A) female (n=42) and male (n=28) or B) by age at time of blood sampling, of those subjects 14-days post first COVID-19 symptoms. Red line marks the mean. **C-D**) IgG OD signals of all subjects were plotted by C) severity of symptoms or D) over time since day of first symptoms. **E-F**) Hospital patients receiving E) immunomodulatory medication (orange, n=31) or F) antiviral medication (purple, n=12) who tested SARS-CoV-2 PCR-positive were assessed for IgG antibodies and compared with pre-COVID-19 controls (open symbols). Statistical analysis was performed using Kruskal-Wallis test using Graphpad Prism software. * p<0.05, ** p<0.01, *** p<0.001.

Within the hospital patient cohort, two groups were of special interest. Those on immunosuppressive therapy and those, receiving anti-viral medication due to infections with either human immunodeficiency virus (HIV) or hepatitis B virus (HBV) (Table 4). Within the patient cohort receiving immunosupressive medication, 7/29 (24%) were asymptomatic for COVID-19, while 3/12 (25%) of those on anti-viral medication did not have COVID-19 symptoms. The average seroconversion rate in both groups was below those seen in the collective patient cohort or the healthcare workers, at 9/29 (31 %) and 9/12 (75%) of those on immunosuppressive medication or anti-viral therapy (Table 4).

**Table 3.**
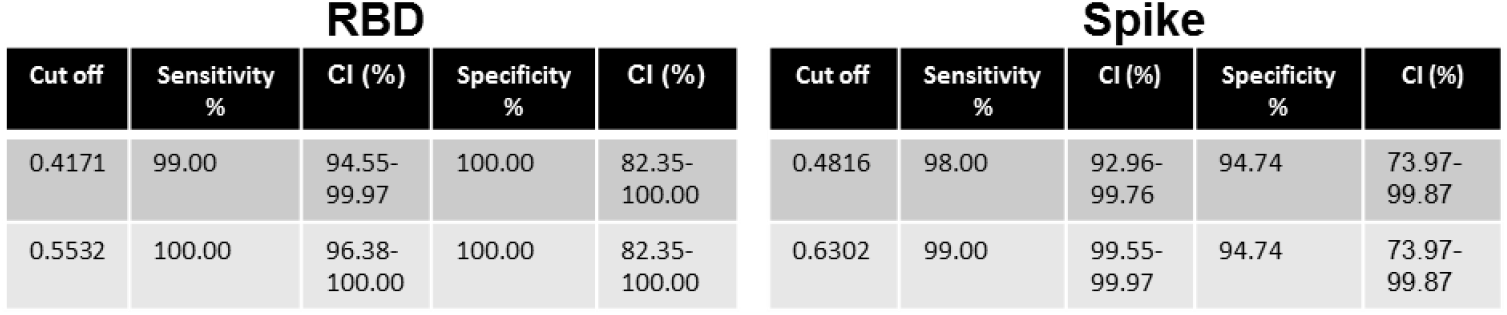
ROC analysis of RBD or Spike protein after initial screening of pre-COVID-19 serum and sera from healthcare workers who were SARS-COV-2 PCR-positive.

**Table 4.**
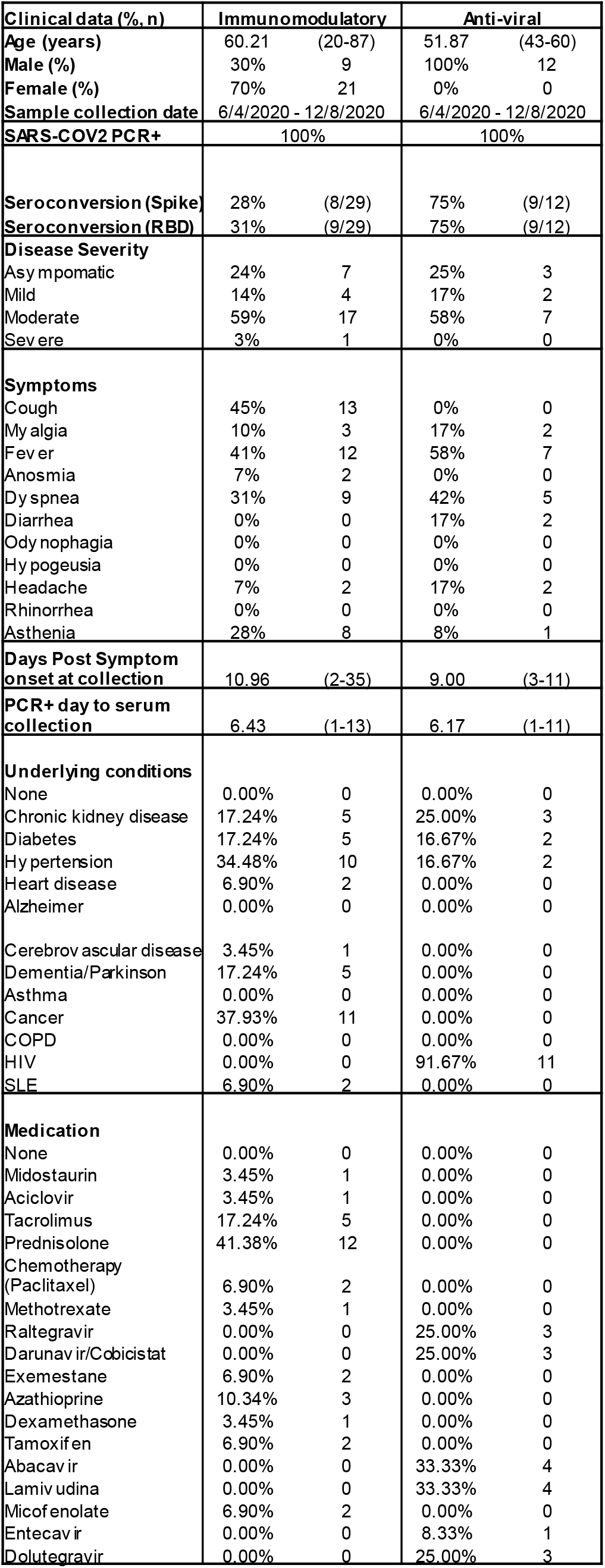
Demographics of patient participants under immunomodulatory or antiviral medication, disease severity categories and symptoms, underlying conditions and medication.

Within the hospital cohort we obtained 29 sera from patients who received immunosuppressive drugs, including 13/29 (40%) receiving the glucocorticoids Prednisolone or Dexamethasone. Sera of these patients were collected between day 2-35 post-COVID-19 symptom onset. In this cohort, all patients tested SARS-CoV-2 positive by PCR, 20/29 (69%) did not seroconvert (Fig.4E). Out of these 20, seven (35%) were asymptomatic, three (15%) were tested within seven days of symptoms. In line with required T cell help for isotype switching, 3/5 (67%) patients on the calcineurin inhibitor Tacrolimus, which was combined with Prednisolone, did not seroconvert. Use of corticosteroids had an inhibitory effect on antibody production, with those patients that seroconverted showing a low signal. The patient on chemotherapy (Paclitaxel) did not show an anti-SARS-CoV-2 RBD IgG response. Three (3/29, 10%) patients receiving immunomodulatory drugs showed a similar anti-SARS-CoV-2 IgG response compared with patients without such medication (day 5-13). Two of these received either Prednisolone or Methotrexate. Although the number of patients tested was modest, these findings indicate that immunosuppresive medications inhibits seroconversion upon SARS-CoV-2 infection.

Twelve samples were from patients who previously contracted HIV1 or HIV2 and were treated with anti-retroviral therapy (Raltegravir, Darunavir/Cobicistat, Lamivudine and Dolutegravir) and one with HBV receiving Entecavir. All patients were male and SARS-CoV-2 seroconversion was analysed within 11 days of symptom onset. Most patients (9/12, 75%) seroconverted (Fig.4F). Of the remaining three HIV patients, two were asymptomatic and one was an early sample taken only three days after COVID-19 symptoms. These results suggest that anti-retroviral medication does not interfere with SARS-CoV-2 seroconversion.

### Large seroconversion screen shows limited SARS-CoV-2 exposure

The University of Lisbon decided to close for all non-essential work early during COVID-19 outbreak, starting from midnight March 13^th^. Since the initial outbreak, reported infection levels in Portugal have remained modest compared with nearby European countries, with 5200 cases per million of the population reported (Johns Hopkins, Worldometer, August 2020). To determine the seroprevalence in University staff, we screened 2571 employees, across all divisions, with serum obtained between May 13^th^ and July 10^th^ 2020. As mentioned, our assay is based on Stadlbauer *et al*., [12] that received emergency FDA approval utilising a two-step method. With the expectation of a low infection prevalence, we first screened staff in a single dilution using the setup as depicted in Figure 5A. We detected 68 samples with an OD above the cut off value of 0.41 (2.6%) (Fig.5B). These samples were subsequently reassessed, using both RBD and Spike protein as well as two serum dilutions, 1/50 and 1/150, as depicted in Figure 5C. Of the 68 tested samples, 38 (56%) were confirmed positive during the second assay (Fig.5D,E), resulting in an infection prevalence of 1.5%. Samples with an intermediate signal for RBD, often just above the cut off, frequently failed the second assay on Spike or even RBD (Fig.5E,F) and some samples with a robust RBD signal did not respond to Spike protein at all. As previously observed (Figure 2), signals between RBD and Spike protein were often comparable, with only a few samples showing a stronger response against Spike. Samples providing a robust signal for RBD often responded similarly with Spike (Fig.5G). To ensure ODs between plates are comparable, the assays inter-plate variation was determined using two dilutions of a QC serum sample or monoclonal antibody (QC_hi_ and QC_lo_) run in each plate. Although day-to-day plate variability is present, this is of very modest amplitude (Fig. 5H). The average and standard deviation of QC_hi_ OD values for the first 12 plates performed were calculated and taken into consideration to validate the following diagnostic plates. The same was done for QC_lo_ values.

**Figure 5.**
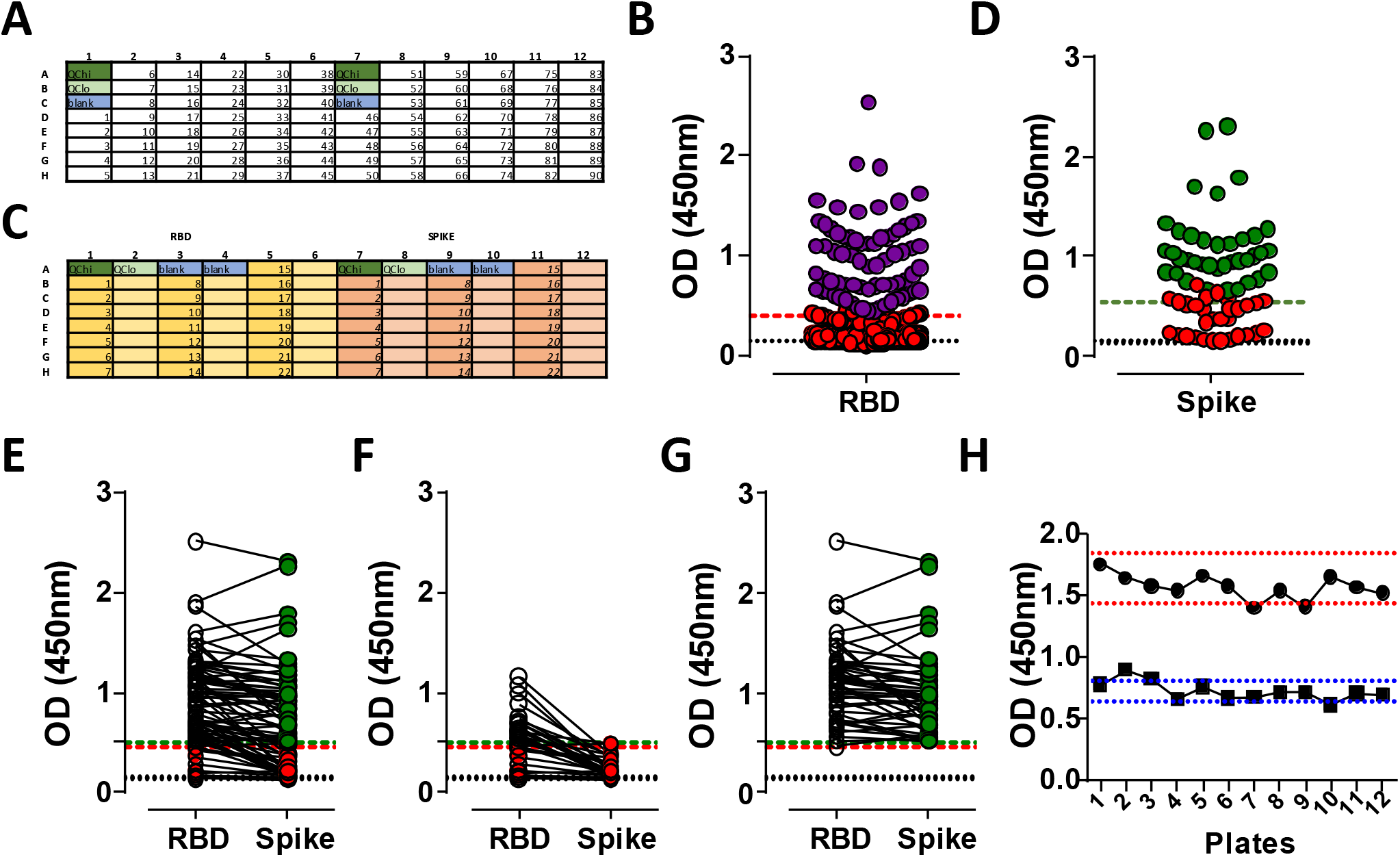
Largescale seroconversion testing of Lisbon University staff. SARS-CoV-2 IgG antibody detection in 1/50 diluted serum samples from University of Lisbon staff using Immulon 4HBX 96-well plates. Absorbance was evaluated at 450 nm. **A**) Schedule of screening plates, coated with 2 μg/ml RBD, accommodating 90 samples/plate, and includes two blanks, two quality control at high concentration (QC_hi_) and two at low concentration (QC_lo_). **B**) Overview of tested staff from Lisbon University (n=2571). Red symbols indicate negative scores, purple symbols indicate ODs above the cut off. **C**) Schedule of re-screening plates, coated with 2 μg/ml RBD (left) or Spike (right), accommodating 21 samples/plate, and includes two dilutions per sample (1/50 and 1/150), two blanks, two QC_hi_ and two QC_lo_ per protein used. **D**) Screening results from the re-screen (n=68), showing samples tested on Spike protein. Green depicts those samples above the cut off for RBD and Spike at both 1/50 and 1/150 dilution (38); Red indicates those samples below the cut off on the second screen for either RBD or Spike (30). **E-G**) Showing RBD and Spike protein signals for the re-assessed samples and an additional 10 negative samples, E) all samples, F) samples assessed negative (open to red symbols) and an additional 10 that were originally negative (red to red symbols), G) samples assessed as positive (open to green symbols). **H**) Quality control signals for 12 sequential plates, showing QC_hi_ (circles) and QC_lo_ (squares). Dotted lines indicate average signal +/- SD for QC_hi_ (red, 200ng/ml) and QC_lo_ (blue (10 ng/ml)) of human anti-SARS-CoV-2.

### Antibody titres follow a classic immune response pattern

To determine antibody responses accurately, we performed serum titrations using RBD protein to assay the anti-SARS-CoV-2 IgM, IgG and IgA responses. In agreement with earlier results, not all subjects assessed early (pre-day 7) after onset of COVID-19 or those asymptomatic show seroconversion, but anti-RBD antibodies rise swiftly during the first days of infection. This is the case for all three isotypes assessed. As with many reported antibody responses, including SARS [15], the anti-SARS-CoV-2 response follows a classic pattern with high antibody responses at the start of the immune response (Suppl.Fig.3A-C).

In addition to early responses from healthcare workers and hospitalised patients, we analysed 187 potential plasma donors for convalescent plasma therapy via the Portuguese Blood and Transplantation Institute (IPST). The volunteers were predominantly male (69%) and on average 38 years old. All were reported to be SARS-CoV-2 PCR positive, on average 100 days prior to serum sample collection (Table 5). COVID-19 symptoms varied from asymptomatic to mild and moderate. At the time of collection, 164/187 (88%) of the potential plasma donors had readily detectable IgG anti-SARS-CoV-2 RBD antibodies. In line with a characteristic immune response, antibody titres assessed in volunteers were reduced compared with titres found during the early immune response in COVID19 patients and healthcare workers, especially IgM and IgA (Suppl.Fig.3D-F).

**Table 5.** Demographics of potential plasma donor participants, disease severity categories (WHO classification), and determined IgG titres.

| **Potential plasma donors** |  |  |
| --- | --- | --- |
| **Age (years)** | 37.92 | (18-58) |
| **Male (%)** | 69% | 128 |
| **Female (%)** | 31% | 57 |
| **Sample collection date** | 8/6/2020 - 14/8/2020 |  |
| **SARS-COV2 PCR+** | 100% |  |
| **Seroconversion (RBD)** | 88% | (164/187) |
| **PCR+ day to serum collection** | 103 | (47-148) |
| **PCR- day to serum collection** | 78 | (34-115) |
| **Titre IgG** | **n** | **%** |
| No titre | 22 | 12% |
| Low (50-300) | 24 | 13% |
| Medium (300-900) | 124 | 66% |
| High (>900) | 17 | 9% |

Analysis of the IgM, total IgG and IgA responses confirmed a rapid and near simultaneous response of the three isotypes tested during the first weeks of SARS-CoV-2 infection (Fig.6A-C). Antibody responses peaked around three weeks after first symptoms, after which the circulating antibody levels reduced. IgM, IgG and IgA peaked at days 15-21 with geometric antibody titre means of 1915, 10695 and 5212 respectively. F rom the second month after disease onset IgG and IgA antibody levels remained readily detectable in most people up to 5 months after first symptoms (Fig.6A-C, Suppl.Fig.3G). Characteristically for an antibody response, IgM titres were low (≤1/200) in 116/163 (71%) of IgG-positive potential plasma donors (Fig.6A-C). Geometric means of IgM, IgG and IgA titres at day 91-120 were 96, 533 and 141, respectively. Early anti-SARS-CoV-2 RBD antibody levels (day 40) were higher in men, with significantly higher titres for all three antibody isotypes, but at late time points (Day 40-150) no differences between men and women were observed (Fig.6D-I). The increase antibody level observed in men was not explained by the severity of COVID-19, with the overall increase in antibodies observed independently of disease symptoms (Suppl.Fig.3H). Furthermore, stratifying subjects within the first 40 days after COVID-19 by severity of symptoms highlighted that increased severity correlates well with increased antibody titres at early stages (Fig.6J-L).

**Figure 6.**
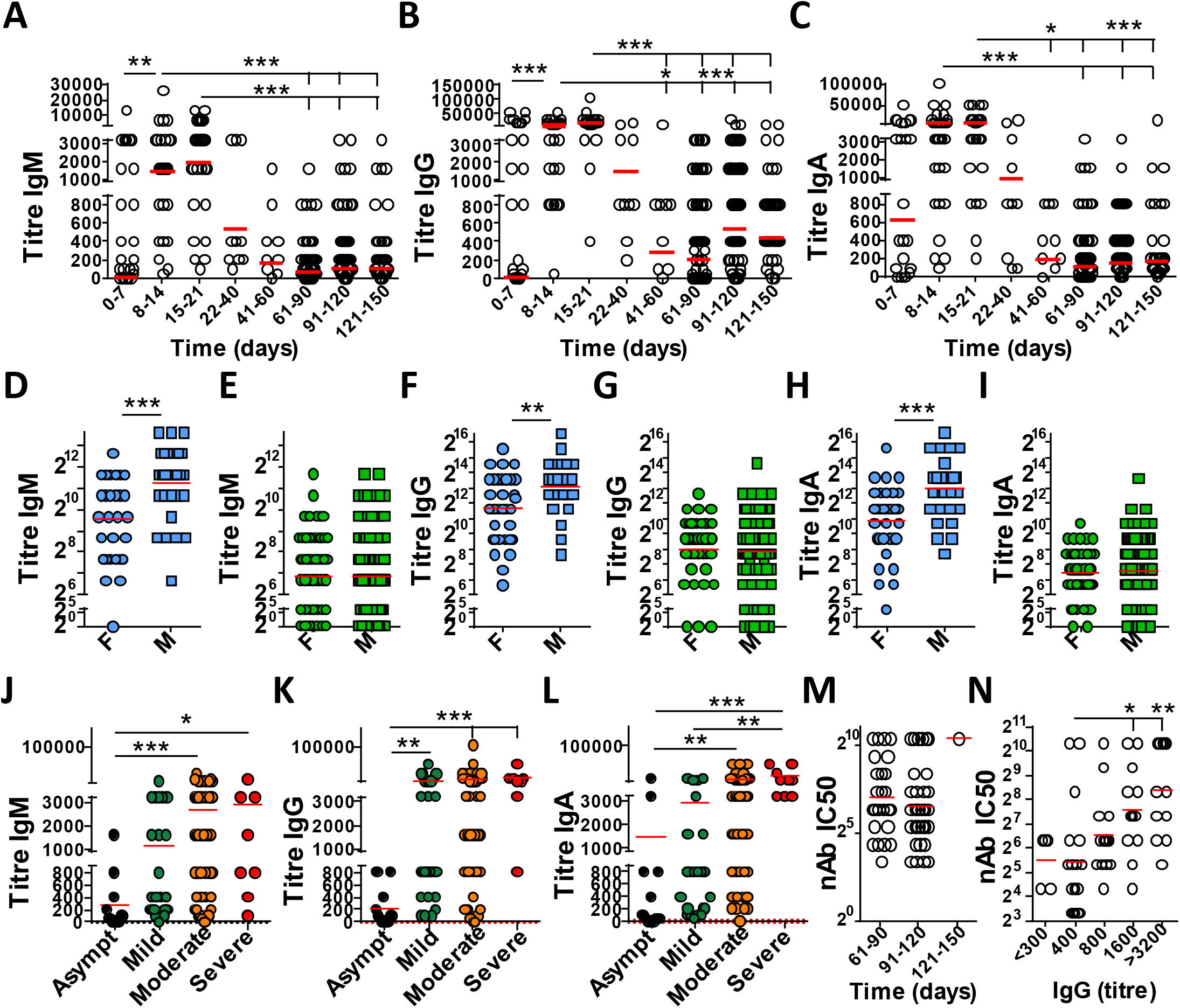
Longitudinal SARS-CoV-2 antibody titres. SARS-CoV-2 antibodies were assessed in 1/50 diluted serum samples from donors in Portugal using Immulon 4HBX 96-well plates. Absorbance was evaluated at 450nm. **A-C**) Anti-SARS-CoV-2 RBD antibody titres plotted over time for A) IgM, B) IgG and C) IgA (n=271). Red line marks the geometric mean. **D-I**) Anti-SARS-CoV-2 antibody titres for males and females during D,F,H) early, days 7-40 (females n=32, males n=29) or E,G,H) late response, days 40-150 (females n=60, males n=114) for D,E) IgM; F,G) IgG or H,I) IgA. Red line marks the mean. **J-L**) Anti-SARS-CoV-2 RBD antibody titres plotted by severity of COVID-19 symptoms experienced for J) IgM, K) IgG and L) IgA. Red line indicates the mean. **M-N**) SARS-CoV-2 neutralising activity was determined in sera (n=60) and plotted against M) time since SARS-CoV-2 PCR+ or N) IgG titre. Red lines indicate geographic mean. Statistical analysis was performed using Mann-Whitney U test (d-i) or Kruskal-Wallis test using Graphpad Prism software. * p<0.05, ** p<0.01, *** p<0.001.

Since titres two months after COVID19 reduced, we next determined the SARS-CoV-2 neutralisaton activity. We found neutralisation activity in all tested sera in which anti-SARS-CoV-2 IgG was determined, across 2-5 months after initial SARS-CoV-2 PCR-positive testing (Fig.6M). The level of SARS-CoV-2 neutralisation activity was found to be proportional to the anti-SARS-CoV-2 RBD IgG titre determined, but not IgM or IgA (Fig.6N, Suppl.Fig.3I-J). Collectively the data highlight that a sustained level of antibodies circulate in the blood for at least 5 months after COVID19, which show SARS-CoV-2 neutralising activity in line with the level of anti-SARS-CoV-2 RBD IgG titres.

## Discussion

For many pathogens and vaccines, antibody titres have been established over the past decades, with accumulating detailed knowledge of average antibody responses. However, every microorganism is different and level, neutralisation activity and longevity of antibody responses can be different between pathogens, vaccines as well as between individuals. Therefore, it remains important to acquire data for each novel infection, especially those posing a threat to human health such as SARS-CoV-2. In addition, initial antibody levels after vaccination, generally correlate well with a significantly reduced risk of (re-)infection and pathology. Thus, the global collection of data from many different geographic locations, patient cohorts and under different local conditions will contribute to a holistic understanding of the new pandemic.

We explain the setup of an ELISA system in detail, as described previously [9], to facilitate implementation in other places and comparison with published results. Although the Spike protein used in the assay is not the only immune-reactive SARS-CoV-2 protein, it has selected because provides additional correlative insights with respect to potential neutralising antibodies present [3, 4, 16]. The assay was setup using samples with high, medium and low titres and an OD of 2.0 was selected to avoid saturating signals. Although serum samples for the assay setup were determined with the initial assays prior to full optimisation, this did not affect their subsequent use.

We show that neither frequently used methods of viral inactivation nor the complexity of RBD, monomeric, dimeric or a mixture of both affects antibody determination. Although reported presence of SARS-CoV-2 is limited in blood [17], the inactivation of serum contributes to reduced risk handling human material. Using the whole RBD fraction from mammalian expression systems will reduce costs due to its superior yield. The higher levels of expression using the relatively small RBD, compared with the full-length Spike protein, makes the use of RBD more economical. We show that individual samples can show differences in signal for either RBD or Spike and that some limited cross reactivity is observed with both proteins. However, there is no disadvantage using RBD compared with the full-length Spike protein, resulting in high specificity and sensitivity. Care should be taken when using serum samples from patients with increased antibody levels. These can be observed in allergies and autoimmune conditions, which may increase the background signal compared with otherwise healthy controls. Most subjects who encountered SARS-CoV-2 seroconverted, although some showed delayed kinetics. Only a few patients did not show an IgG response that was not explained by early sampling (<day 7) or an underlying condition that required the use of immunosuppressive drugs. Most of the non-responders were asymptomatic, which may point to a very modest immune response upon encountering a low viral load, or, since SARS-CoV-2 PCRs also generates false-positives, these participants may not have been infected.

Within the patient cohort there were additional groups of interest. With SARS-CoV-2 being a positive stranded RNA virus, the use of anti-retroviral medication had no major effect on antibody responses against the virus. The mode of action of these drugs are not known to interfere with RNA viruses or antibody production. Our data show the successful management of the eleven patients previously infected with HIV, with all of those patients showing a robust SARS-CoV-2 antibody response after the second week of COVID-19 symptoms, in line with previous works [18, 19]. The use of immunomodulatory drugs had an inhibitory effect on SARS-CoV-2 seroconversion. This is in line with the mode of action of these drugs, inhibiting the activation or production of lymphocytes. Although this will inhibit the clearance of SARS-CoV-2, the use of these immune-inhibitory drugs such as Dexamethasone could be beneficial in cases of severe immune response against SARS-CoV-2, resulting in cytokine storm and immunopathology [20].

We and others [4] found higher antibody titres in men compared with women. This is surprising since women on average have more B cells and produce more antibodies [21]. Higher antibody titres in men, only observed during the acute stage, correlates well with men showing more severe symptoms and increased fatality, as reported [22, 23]. Innate antiviral responses, such as those mediated via toll-like receptor-7, are enhanced in women [24], which may explain their increased resistance against SARS-CoV-2, similarly to influenza virus [25].

In many countries implementing mitigation strategies, infection prevalence remained modest at the time of sampling, May-June 2020, with low frequency of infection [26-29]. This increases the proportional contribution of any false positives to the result. The ELISA assay as used, was approved by the FDA as a two-step method (https://www.fda.gov/media/137029/download). We show that using only one protein for a large population screen picks up some false positives (30/2571; 1.2%). Especially when the infection rate is low, the two-step method is highly beneficial and strongly recommended [27]. Furthermore, the introduction of an additional dilution step ensures robustness, resulting in 1.5% (38/2571) of the staff members that seroconverted. Overall, there were limited differences between RBD and Spike reactivity, with a few samples responding more robustly to Spike, possibly reflecting the increased amount of epitopes available in the larger protein.

The question of long-lasting and protective immunity against SARS-CoV-2 is a focus of current research. We show that the initial antibody response against SARS-CoV-2 raises the three main isotypes in close concert as previously reported for SARS as well as SARS-CoV-2 [15, 30]. The kinetics of the response follows a well-known pattern with antibody levels peaking around 3 weeks after symptoms and declining thereafter. Late responses are characterised by low or sometimes undetectable levels of IgM, modest IgA, but at least until 150 days post-PCR-positive reaction, mostly a robust IgG response. Between days 40-150 we found 89% of previous SARS-CoV-2-PCR-positive subjects (177/199), healthcare workers and potential plasma donors, to carry antibodies, 75% of which had medium to high titres (>300). In addition, we found that in subjects with detectable anti-SARS-CoV-2 IgG, neutralisation activity was in accordance with the determined IgG titre level. This is in agreement with a recent report [16]. This and the strong correlation between RBD IgG titres and neutralising activity as well as protective immunity [16, 31], suggests that most people infected with SARS-CoV-2 will have circulating protective immunity for many months after COVID-19. In addition, recent reports of T cell responsiveness [32-35] show a robust T cell response. Since the SARS-CoV-2 response is in line with well-known and detailed studied immune responses resulting in lymphocyte memory, it is very likely that SARS-CoV-2 protective immunity, reducing disease severity, will last for at least a few years.

## Materials and methods

### SARS-CoV-2 RBD and Spike protein constructs

RBD and Spike protein constructs were obtained from Dr Florian Krammer, Icahn School of Medicine at Mount Sinai, New York, USA.

### Production, purification of antigen recombinant proteins

Production and purification of recombinant proteins was performed at Instituto de Biologia Experimental e Tecnológica (iBET) Oeiras, Portugal as part of the Serology4COVID consortium as previously described by Stadlbauer *et al*., 2020 [12]. Briefly, Spike or RBD antigen containing His-tag is produced by transient transfection of Expi293F™ cells (Thermo Fisher Scientific) with plasmids suitable for mammalian cell expression (pCAGGS), harbouring spike gene or RBD gene, respectively. All purification steps were performed at 4°C. At 3 days post-transfection, cultures are centrifuged, supernatants are collected and filtered through Sartopore MidiCaps. The clarified supernatants are concentrated and dialysed with binding buffer by tangential flow filtration, using 10 kDa or 30 kDa membranes, for RBD or Spike purification, respectively. The final dialysed and concentrated sample is filtered through 0.22 μm membrane and loaded into HisTrap HP columns, equilibrated with binding buffer. Proteins are eluted with a linear gradient up to 500 mM Imidazole. The fractions containing Spike or RBD are concentrated to 1-2 mg/ml using Vivaflow 200 crossflow devices. Removal of imidazole and exchange to PBS buffer is performed by diafiltration with 10 volumes of PBS. Protein concentration is determined by A280nm combined with the specific extinction coefficient. The concentrated and formulated products are filtered through 0.22 jm membrane, aliquoted, snap frozen in liquid nitrogen and stored at −80 °C.

### Human samples collection

Upon informed consent, blood was taken by vein puncture and two BD Vacutainer CPT tubes of blood and one serum tube were collected per patient. For serum collection, tubes were centrifuged at 2200 rpm, 10 min at 4^°^C and upper 6 × 0.25ml of serum placed into six cryotubes. Samples are stored in a −80^°^C ultra low freezer at the iMM Biobank.

Serum samples were obtained from the iMM biobank COVID-19 collection, and pre-pandemic control sera from two allergy collections. Patients were recruited from the COVID-19 unit and the Allergy Department of Hospital de Santa Maria, Centro Hospitalar Lisboa Norte. The COVID-19 collection and scientific use was approved by the Lisbon Academic Medical Center Ethics Committee (Ref. n.° 155/20) as was the staff screening (Ref. n.° 181/20). The allergy studies were approved with reference 116/13. Potential plasma donors registered voluntarily via the IPST website. Criteria for registration were a diagnosis of COVID-19 documented by a positive PCR test for SARS-CoV2 followed by two negative or one negative PCR tests 14 or 28 days prior to collection, respectively. Medical interviews were conducted to ensure that the criteria for apheresis plasma donation were fulfilled and that a fully recovery from COVID-19 had been achieved.

Signed informed consent was obtained from all volunteers. All data were treated confidentially and anonymous, according to (EU) 2016/679 of 27 April 2016 (General Data Protection Regulation). A professional obliged to confidentiality with guarantee appropriate information security measures carried out the data collection under the terms of the GDPR paragraph 2, article 29 Law no. 58 /2019, 8^th^ August.

### Virus inactivation

To reduce risk from any potential residual virus in the serum, three different virus inactivation methods were tested: incubation for one hour at 56°C (heat inactivation), addition of 0.1% Triton X-100 (a non-ionic surfactant), or the combination of both (H+ Tx).

### Antibody measurements

Anti-SARS-CoV-2 ELISAs were performed as described previously [12]. Briefly, flat bottom 96-well plates (Immulon 4 HBX Thermo Scientific) were coated with recombinant protein RBD or Spike prepared in PBS at a concentration of 2 μg/ml (50 μl/weN) overnight at 4°C. Coated plates were washed with PBS-0.05%Tween (PBS-T) using a Well-wash 1×8 (Thermo Fisher Scientific) or Aquamax200, 3x for IgG detection and 10x for IgM analysis. Plates were blocked with 200 μl/well of 3% non-fat milk powder in PBS-1%T for 1 hour at room temperature and then washed with PBS-T 3x or 10x, as described previously. Serum samples were diluted in PBS-0.1%T + 1% non-fat milk powder, added (100 μl/well) and incubated for 1-2 hours at room temperature, washed with PBS-T 3x or 10x Hereafter several antibody isotypes, namely Total Ig, IgG, IgM and IgA anti-SARS-CoV2 were detected using horseradish peroxidase (HRP)-labelled goat anti-human IgG+IgM+IgA (Abcam, ab102420), IgG Fc (Abcam, ab97225), IgM mu chain (Abcam, ab97205), IgA alpha chain (Abcam, ab97215) respectively. Secondary antibodies were diluted in PBS-0.1%T + 1% non-fat milk powder (50 μl/well) and added for 1 hour at room temperature, washed with PBS-T 3x or 10x, and developed with TMB substrate solution (TMB Substrate Reagent Set, BD OptEIA™, 555214), 100μl/well for 10 minutes. The reaction was stopped with 1M sulphuric acid (50 μl/well) and optical density at 450nm was measured via Infinite M200 (TECAN) plate reader. Each plate contained a Quality control (QC) sample, composed of a pool of positive samples or monoclonal antibody, tested in a high and low dilution. For material details, see methods supplement.

### Neutralisation assays

Anti-SARS-CoV-2 ELISAs were performed as described in detail recently [36]. SARS-CoV-2 pseudo particles (pp) were a kindly provided by Dr Benhur Lee, Icahn School of Medicine at Mount Sinai, New York, USA. Briefly, 24 hours prior to infection, Vero CCL81 cells grown in DMEM supplemented with 10% FBS were seeded at 20,000 per well in a 96-well plate. SARS-CoV-2pp and serial dilutions of sera (1/10 in DMEM with 10% FBS, and further 2-fold dilutions) were incubated at room temperature for 30 minutes. Media from Vero cells was substituted with the SARS-CoV-2pp/serum mix; plates were spinoculated by centrifugation at 1250rpm for 1 hr at 37°C. After overnight incubation at 37°C, culture medium was removed, and Renilla luciferase production was assessed on Tecan 2 plate reader using the Dual-luciferase Reporter Assay system (Promega), according to manufacturer instructions. IC50 were determined as the last serum dilution at which the titration curve matches inhibition equal or above of 50% of the 100% assay.

### Statistical analyses

A Kruskal-Wallis test (non-parametric test) was done to compare the geometric ratios between groups with a significance level of 0.05 (Dunn’s multiple comparisons test), student t test or two-way ANOVA were used as stated in the figure legends, calculated using GraphPad Prism 6.0 software.

## Data Availability

Supplementary methods and figures have been submitted. Any data will be made available upon reasonable request. This manuscript does not contain other large data sets, but for ELISA plate readouts.

## Acknowledgements

We would like to thank all volunteers who helped with blood collections (AR Pires, A Ramalho-dos-Santos, A Biscaia-Santos, F Ribeiro, S Caetano, P Napoleão, MJ Silva, P Alves, R Pedroso, P Corredeira, A Friães, M de Niz, I Bento, S Pereira, S Mensurado) and donors and patients for providing blood samples and cooperation to make this study possible. Serum samples were requested from Biobanco-IMM, Lisbon Academic Medical Centre, Lisbon, Portugal.

We like to acknowledge the funding from the European Union H2020 ERA project (No 667824 - EXCELLtoINNOV) and the Fundação para a Ciência e a Tecnologia (FCT) to PF-C. (SFRH/BD/131605/2017), PTDC/MED-IMU/28003/2017, and research4COVID19 (n° 231_596873172, Generating SARS-CoV2 seroconversion assay and n° 729, High-throughput SARS-CoV2 neutralising antibodies assessment), with additional support by Sociedade Francisco Manuel dos Santos.

We would like to acknowledge the generous sharing of the expression constructs by Drs Florian Krammer and Benhur Lee, Icahn School of Medicine at Mount Sinai, New York, USA (Development of SARS-CoV-2 reagents was partially supported by the NIAID Centers of Excellence for Influenza Research and Surveillance (CEIRS) contract HHSN272201400008C), and the protein production by Drs Paula Alves, Pedro Cruz and Rute Castro at Instituto de Biologia Experimental e Tecnológica (iBET) Oeiras, Portugal as part of the Serology4COVID consortium.

